# An Epidemic Model for Multi-Intervention Outbreaks

**DOI:** 10.1101/2023.06.27.23291973

**Authors:** Kathryn L. Schaber, Sagar Kumar, Baker Lubwama, Angel Desai, Maimuna S. Majumder

## Abstract

Modeling is an important tool to utilize at the beginning of an infectious disease outbreak, as it allows estimation of parameters—such as the basic reproduction number, R_0_—that can be used to postulate how the outbreak may continue to spread. However, there exist many challenges that need to be accounted for, such as an unknown first case date, retrospective reporting of ‘probable’ cases, changing dynamics between case count and death count trends, and the implementation of multiple control efforts and their delayed or diminished effects. Using the near-daily data provided from the recent outbreak of Sudan ebolavirus in Uganda as a case study, we create a model and present a framework aimed at overcoming these aforementioned challenges. The impact of each challenge is examined by comparing model estimates and fits throughout our framework. Indeed, we found that allowing for multiple fatality rates over the course of an outbreak generally resulted in better fitting models. On the other hand, not knowing the start date of an outbreak appeared to have large and non-uniform effects on parameter estimates, particularly at the beginning stages of an outbreak. While models that did not account for the decaying effect of interventions on transmission underestimated R_0_, all decay models run on the full dataset yielded precise R_0_ estimates, demonstrating the robustness of R_0_ as a measure of disease spread when examining data from the entire outbreak.

## Introduction

Epidemiological modeling is an important tool at the beginning of an infectious disease outbreak, as it allows estimation of parameters that can be used to postulate how the outbreak may continue to spread. By estimating the transmission rate of an infectious agent at the beginning of an outbreak, the basic reproduction number (R_0_) can be calculated. Defined as the number of secondary infections generated by an infected index case in a completely susceptible population, R_0_ is used to determine the epidemic potential of an outbreak, where values above one indicate epidemic potential and values below one typically lead to the end of the outbreak (1–4). This value is particularly important as it is often used to determine which control efforts should be put in place, as seen recently at the beginning of the COVID-19 pandemic (5–8).

However, many challenges exist in estimating accurate reproduction numbers throughout the course of an outbreak, particularly in the case of emerging infectious diseases where little prior knowledge is available. In the early stage, information is rarely available regarding the primary case (i.e., the first case in a given population), thus often leaving the start date of the outbreak unknown (9, 10). Further, once an index case (i.e., the first infected individual reported to health authorities) has been identified, the first few days of data collection tend to yield large increases in case counts due to “probable” cases being identified retrospectively (11). This identification of probable cases can be particularly complex for diseases with high case fatality rates (CFRs), where post-mortem individuals are reported as a case and a death on the same date, closely tying the trends between these values and causing inflated CFR estimates that decrease over the course of the outbreak (12, 13). As the outbreak progresses, improved surveillance, reporting efforts, and public awareness can decrease the time between symptom onset and case reporting ((14, 15). Accounting for these changing dynamics between case count and death count trends over the course of an outbreak is not something that can be easily accounted for when modeling disease spread (16, 17).

The implementation of control efforts during an outbreak, particularly in the early phase, can also add challenges to modeling efforts. While the presence of an intervention can reduce the initial transmission rate, there is no way to estimate the true epidemic potential of the pathogen using R_0_ when control efforts are implemented immediately after detection of the index case, as the intervention would have already changed the effective susceptibility of the population when data reporting started. In this situation, the effective reproduction number (R_e_) is typically used to predict the number of secondary infections generated by an infected index case — namely, in a population that is not completely susceptible (4, 18, 19). R_e_ can be estimated throughout the course of an epidemic to assess changes in disease transmission due to infection-conferred immunity and control efforts, such as social distancing, lockdown, or vaccination (4, 19–21).

There have been disease models created that account for decaying transmission due to control implementation (22–27). However, to our knowledge, these models do not allow for the implementation of multiple control efforts with differing effects on disease spread during the outbreak. Indeed, interventions can not only have differing effects on transmission, but also have varying time delays between implementation and impact, as seen with the COVID-19 pandemic (28–30). Further, not all interventions are sustainable and some may have diminished effects on pathogen transmission (31). For example, contact tracing with a limited number of resources becomes inefficient when case counts increase; similarly, social distancing adherence can decrease over time due to “pandemic fatigue”, as seen with COVID-19 (32–34). There is a need to account for the presence of multiple control efforts, their delayed effects, and the possible diminishing of those effects at the beginning of an outbreak to determine intervention efficacy.

Recently, this need was underscored when an outbreak of Ebola disease caused by Sudan virus (SUDV), recently reclassified as Sudan Virus Disease (SVD) (35, 36), occurred in Uganda for the first time in over a decade. As SVD outbreaks have historically been smaller and more sporadic than their Ebola Virus Disease counterparts, there has been considerably less research done on SVD transmission patterns, treatment methods, and vaccine candidates (37). During the recent SVD outbreak in Uganda, near-daily data were reported from detection of the first case to the last case, providing an invaluable case study in overcoming the challenges of modeling an emerging infectious disease outbreak, particularly at the beginning stages.

Here, using the 2022 outbreak of SVD in Uganda as a case study, we create a model and present a workflow which accounts for probable case counts, changing dynamics between case and death count trends, and the presence of multiple control efforts and their delayed or diminished effects. By examining how including, or not including, each of these factors can change estimates of disease spread and epidemic potential, we determine which are the most important to account for in models of future outbreaks.

## Methods

### Model

To examine the trajectory of the outbreak and the effect of two separate control measures, a susceptible-exposed-infectious-deceased-recovered (SEIDR) model was utilized. This is a modified version of the classic SEIR model, previously used to model Ebola disease spread (38), which accounts for disease-related mortality. We chose to include the deceased class (D) in the model due to the high case fatality rates associated with SVD (39). Further, this allows our model to have a closed system, defined by the following differential equations:

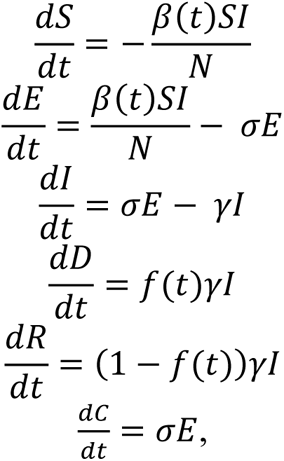

where classes S, E, I, D, and R denote the number of susceptible, exposed, infectious, deceased, and recovered individuals and the total population, N = S + E +I + D + R. C is not an epidemiological state; rather it is used to keep track of the cumulative number of cases over time. The average duration of incubation, which is the time it takes a disease to develop after exposure to a pathogen, is 1/σ; 1/γ is the average duration of infectiousness; f(t) is the case fatality rate (CFR) at time t; and β(t) is the time-dependent transmission rate. To account for the presence of *n* different intervention efforts at separate time points, τ_1_,τ_2_, …, τ_*n*_, β(t) is set as a constant up until the introduction of the first intervention at time τ_1_, at which point it begins to decay exponentially at rate k_1_. The diminished efficacy of interventions is accounted for by allowing the decaying effects of intervention *x* to end at some time point, τ_*x_end*_, after which the transmission rate is set at a new constant value, β_*x*+1_, and the next intervention has its own independent effect on transmission rate decay, *k_x+1_*:

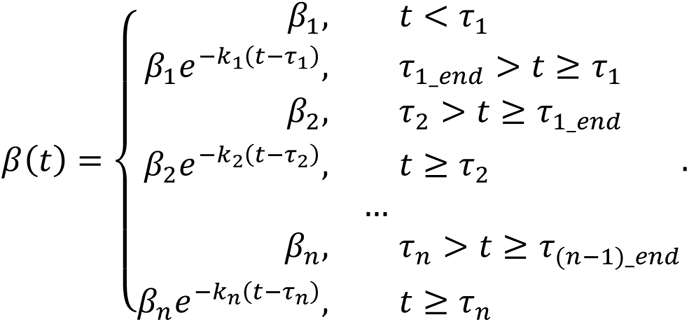

This builds upon previous models of SVD that account for exponentially decaying transmission after a single intervention (22, 23). Changes in CFR may also be seen throughout the course of an outbreak due to a decrease in the time between symptom onset, care seeking, and case detection, potentially allowing for earlier supportive care and improved survival. Further, for diseases with a high CFR, such as SVD, early estimates are likely to be inflated from retrospective case/death reporting. Thus, for each time point of intervention implementation in our model (τ_1_,τ_2_, …, τ_*n*_), we allow the option for the CFR, f(t), to change to a new constant value:

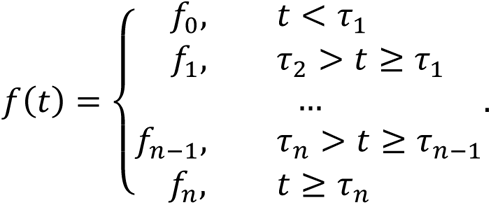

This model will be henceforth referred to as the “independent decay model”.

### SVD Outbreak and Datasets

On September 20th, 2022, for the first time in over a decade, Uganda declared an outbreak of Ebola disease caused by Sudan virus (SUDV) following confirmation of a case in Mubende district (40). Sudan Virus Disease (SVD) is a severe, often fatal, zoonotic illness that spreads through contact with the bodily fluids of a person who is sick with or has died from the disease (39, 41–43). Unlike with Ebola virus, there are currently no treatments or vaccines targeting SUDV, although there were a few vaccine candidates that became prioritized and began clinical trials in Uganda during this outbreak (44–47). The last case of the SVD outbreak was reported on November 27th; however, the Ministry of Health of Uganda did not declare the end of the outbreak until January 11th, 2023, 42 days (twice the maximum incubation period of SUDV infections) after the last confirmed case was buried (48).

During this SVD outbreak, 22 of the 164 cases and 77 deaths were reported as “probable”, with the majority (N=19) occurring before the index case was detected (48, 49). Two interventions were enacted over the course of the outbreak. First, a rapid and extensive contact tracing effort was put in place after the outbreak was declared on September 20th, with 4793 cumulative contacts listed and around 91% of contacts completing 21 days of follow-up (although there were some inconsistencies in reports of contacts completing the 21-day follow-up, as cataloged in Text S1) (49, 50). Second, a 21-day lockdown was declared in the two most affected districts (Mubende and Kassanda) on October 15th, which included an overnight curfew, the closure of places of worship and entertainment, and restriction of movements in and out of the districts (50, 51). The lockdown was extended twice, with mobility restrictions finally being lifted 63 days later on December 17th (52). While the majority of cases occurred in Mubende, Kassanda, and Kampala districts, there were nine total districts with confirmed cases: Mubende, Kassanda, Kampala, Kyegegwa, Bunyangabu, Kadagi, Wakiso, Masaka, and Jinja (48).

Our model used multiple sources to procure data on the number of cases and subsequent deaths reported during this SVD outbreak in Uganda. By September 29, 2022, the Ministry of Health (MoH) Uganda began publishing near-daily situation reports on the SVD outbreak, providing data on the number of confirmed and probable cases and deaths (49). For the nine days following the report of the first Ebola case on September 20, 2022, these reports were not yet available. Thus, we instead sourced case and death counts from the MoH Uganda and World Health Organization (WHO) Regional Office for Africa Twitter pages and the WHO Uganda news updates (see Text S2 for date-specific sources) (40, 49, 53–56). Daily case and death counts were compiled in two ways: using only those reported as confirmed (the “confirmed reported only” dataset) and using both confirmed and probable values to obtain daily counts for total cases and deaths (the “as reported” dataset). Together, these datasets covered the period from September 20, 2022 – December 4, 2022. While the last case was reported on November 27th, the last change to case and death counts was made a week later due to data reconciliation efforts (Text S1) (49).

A third dataset (aside from the “confirmed reported only” and “as reported” datasets) was also compiled by extracting information from graphs in the MoH Uganda situation reports, where cases were indexed by the onset date of their symptoms. This dataset provided a timeline for those probable cases that occurred at the beginning of the outbreak that were not reported until days or weeks after death, thus allowing us to examine the time between symptom onset and case reporting throughout the outbreak. This will henceforth be referred to as the “MoH onset” dataset.

The difference between symptom onset and case reporting dates was examined by plotting the time series of cumulative case counts by symptom onset date (“MoH onset”) shifted zero, two, four, and six days forward alongside the cumulative case counts “as reported”. As the “MoH onset” dataset included both probable and confirmed cases, comparisons were made with the “as reported” dataset rather than the “confirmed cases only”. Mean square error estimates (MSE) were calculated to compare the datasets. The “MoH onset” and the “as reported” datasets were also used to estimate the effective reproduction number (R_e_), with time-varying results compared over the course of the outbreak. R_e_ was estimated using the EpiEstim package in R (57) with the default weekly sliding windows, where values were estimated on the last day of the weeklong window. The serial interval (i.e., the time between symptom onset for two consecutive cases in a chain of transmission) was set to be parametric following a Weibull distribution with mean of 12.0 days and standard deviation of 5.2 days. This was based on contact tracing data from the 2000 SVD outbreak in Uganda (38). Time-varying R_e_ estimates from both datasets were graphed for comparison. To examine the changes in the case fatality rate (CFR) throughout the outbreak due to decreased case detection time, daily CFRs were calculated. Further, to determine the effect of retrospective “probable” death reporting early in the outbreak, daily CFRs were calculated for both the “as reported” dataset and the “confirmed reported only” dataset. A daily Chi-square Test of Homogeneity with Yates’ continuity correction was conducted to determine whether the proportion of deaths (CFR) was equivalent when considering only confirmed cases (“confirmed reported only”) as when considering confirmed and probable cases (“as reported”). These values were also graphed for visual comparison.

### Parameter Estimation Workflow

For all models, duration of incubation (1/σ) and infectiousness (1/γ) were set at 3.35 and 3.5 days, respectively, based on previous estimates from the 2000 SVD outbreak in Uganda (38, 58). For all model parameterizations, maximum likelihood estimates (MLE) of the free parameters were obtained by fitting models to case and death count data using the bbmle package in R and the Nelder & Mead optimization algorithm (59, 60). Cumulative case and death counts were assumed to be Poisson distributed. All models were fit to the “as reported” case and death count dataset, because reported data are generally what is available when conducting early outbreak modeling. Using the MLE values for initial transmission rate (β_1_) and the fixed value for duration of infectiousness (1/γ), the basic reproduction number (R_0_) was calculated as R0 = β_1_/γ. To determine how accurately each model parameterization fit to the reported data, MLE values were plugged back into the SEIDR model and simulations provided the predicted daily incidence counts and cumulative case and death counts over time. These predicted values were then compared to the observed “as reported” dataset using MSE. The predicted cumulative case counts were also used for Pearson goodness-of-fit tests, examining how well the observed case counts corresponded to those predicted from each model.

Model parameterization followed a branching process, as depicted in Figure S1. This workflow was utilized in order to examine the individual and combined effects of accounting for probable case counts, changing dynamics between case and death count trends, and the presence of multiple control efforts and their delayed or diminished effects.

#### 1. First intervention

##### a. Decaying transmission

To examine the effect of the first intervention — the extensive contact tracing effort — on transmission (n=1), the model was parameterized using only data from September 20^th^ – October 14^th^, before the second intervention — the 21-day lockdown — was enacted. The model was parameterized in two ways — as a base SEIDR model where the intervention had no effect (k_1_=0):

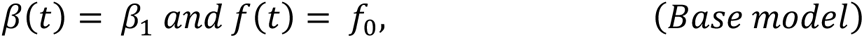

and as a model like Althaus et al. (23), where transmission decayed after the intervention was implemented:

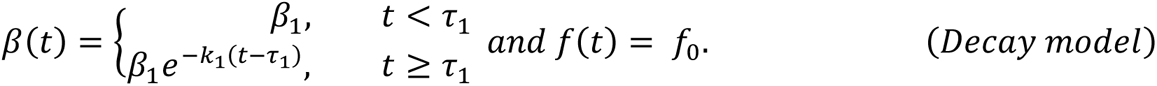

For both scenarios, as in Althaus et. al. (23), we also accounted for the outbreak start date (i.e., the date of the primary case’s symptom onset, τ_0_) in two ways: (1) it was fixed as August 7^th^, 2022, based on data from MoH Uganda and the Centers for Disease Control and Prevention (CDC) and (2) it was allowed to be a free parameter estimated by maximum likelihood estimation (MLE) (Table 1) (61). The latter option was included to reflect the state of knowledge early on in an outbreak when the date of symptom onset for the primary case is typically not yet known (10).

**Table 1:** List of models run.

| Step | | Data range | | Model Type | Days delayed | | Primary case date ($\tau_0$) fixed? | CFR change at start of: | |
| --- | --- | --- | --- | --- | --- | --- | --- | --- | --- |
|  |  | Start date | End date |  | 1<sup>st</sup> intervention | 2<sup>nd</sup> intervention |  | 1<sup>st</sup> intervention? | 2<sup>nd</sup> intervention? |
| 1a | i | 9/20/22 | 10/14/22 | Base | 0 | --- | N | N | --- |
| 1a | ii | 9/20/22 | 10/14/22 | Base | 0 | --- | Y | N | --- |
| 1a | iii | 9/20/22 | 10/14/22 | Decay | 0 | --- | N | N | --- |
| 1a | iv | 9/20/22 | 10/14/22 | Decay | 0 | --- | Y | N | --- |
| 1b | i | 9/24/22 | 10/14/22 | Base | 0 | --- | N | N | --- |
| 1b | ii | 9/24/22 | 10/14/22 | Base | 0 | --- | Y | N | --- |
| 1b | iii | 9/24/22 | 10/14/22 | Decay | 0 | --- | N | N | --- |
| 1b | iv | 9/24/22 | 10/14/22 | Decay | 0 | --- | Y | N | --- |
| 1c | i | 9/24/22 | 10/14/22 | Decay | 4 | --- | N | N | --- |
| 1c | ii | 9/24/22 | 10/14/22 | Decay | 4 | --- | Y | N | --- |
| 1c | iii | 9/24/22 | 10/14/22 | Decay | 7 | --- | N | N | --- |
| 1c | iv | 9/24/22 | 10/14/22 | Decay | 7 | --- | Y | N | --- |
| 1c | v | 9/24/22 | 10/14/22 | Decay | 10 | --- | N | N | --- |
| 1c | vi | 9/24/22 | 10/14/22 | Decay | 10 | --- | Y | N | --- |
| 1d | i | 9/24/22 | 10/14/22 | Decay, CFR change | 0 | --- | N | Y | --- |
| 1d | ii | 9/24/22 | 10/14/22 | Decay, CFR change | 0 | --- | Y | Y | --- |
| 1d | iii | 9/24/22 | 10/14/22 | Decay, CFR change | 4 | --- | N | Y | --- |
| 1d | iv | 9/24/22 | 10/14/22 | Decay, CFR change | 4 | --- | Y | Y | --- |
| 1d | v | 9/24/22 | 10/14/22 | Decay, CFR change | 7 | --- | N | Y | --- |
| 1d | vi | 9/24/22 | 10/14/22 | Decay, CFR change | 7 | --- | Y | Y | --- |
| 1d | vii | 9/24/22 | 10/14/22 | Decay, CFR change | 10 | --- | N | Y | --- |
| 1d | viii | 9/24/22 | 10/14/22 | Decay, CFR change | 10 | --- | Y | Y | --- |
| 2a | i | 9/24/22 | 12/4/22 | Decay | 7 (above) | --- | Y | N (above) | --- |
| 2a | ii | 9/24/22 | 12/4/22 | Two decay | 7 (above) | 0 | Y | N (above) | N |
| 2b | i | 9/24/22 | 12/4/22 | Two decay | 7 (above) | 4 | Y | N (above) | N |
| 2b | ii | 9/24/22 | 12/4/22 | Two decay | 7 (above) | 7 | Y | N (above) | N |
| 2b | iii | 9/24/22 | 12/4/22 | Two decay | 7 (above) | 10 | Y | N (above) | N |
| 2c | i | 9/24/22 | 12/4/22 | Two decay, CFR change | 7 (above) | 0 | Y | N (above) | Y |
| 2c | ii | 9/24/22 | 12/4/22 | Two decay, CFR change | 7 (above) | 4 | Y | N (above) | Y |
| 2c | iii | 9/24/22 | 12/4/22 | Two decay, CFR change | 7 (above) | 7 | Y | N (above) | Y |
| 2c | iv | 9/24/22 | 12/4/22 | Two decay, CFR change | 7 (above) | 10 | Y | N (above) | Y |
| 3a | i | 9/24/22 | 12/4/22 | Two decay, diminished fx | 7 (above) | 4 | Y | N (above) | N |
| 3a | ii | 9/24/22 | 12/4/22 | Two decay, diminished fx | 7 (above) | 7 | Y | N (above) | N |
| 3a | iii | 9/24/22 | 12/4/22 | Two decay, diminished fx | 7 (above) | 10 | Y | N (above) | N |
| 3b | i | 9/24/22 | 12/4/22 | Two decay, diminished fx, CFR change | 7 (above) | 4 | Y | N (above) | Y |
| 3b | ii | 9/24/22 | 12/4/22 | Two decay, diminished fx, CFR change | 7 (above) | 7 | Y | N (above) | Y |
| 3b | iii | 9/24/22 | 12/4/22 | Two decay, diminished fx, CFR change | 7 (above) | 10 | Y | N (above) | Y |

##### b. Onboarding of data collection

The “onboarding” period of data collection refers to the first few days after the identification of an outbreak, when there are large increases in case and death counts due to “probable” cases being identified retrospectively and/or post-mortem. To examine the effect of excluding these first few days of data collection (September 20th – 23rd) when estimating disease spread parameters, the four parameterizations from above (i.e., base and decay models with the outbreak start date fixed and free) were also run for data from September 24^th^ – October 14^th^. MSE values for September 24^th^ – October 14^th^ were compared for the resulting eight parameterizations to determine if inclusion or exclusion of the data onboarding period yielded daily incidence counts and cumulative case and death counts more similar to those from the reported dataset.

##### c. Delayed decay

For the better fitting timeframe (September 20th – October 14th vs. September 24th – October 14th), we also parameterized the model to delay the effect of intervention by 4, 7, and 10 days by setting τ_1_to be September 24^th^,27^th^, and 30^th^, respectively. This was done to account for the fact that the effects of control measures cannot typically be observed immediately after implementation. Further, in the case of contact tracing, it may take a few days to acquire the available resources/personnel required and to detect all the infected cases for tracing. These 4-, 7-, and 10-day delay scenarios were run with both free and fixed τ_0_ values (Table 1).

##### d. Changing case fatality rates

Due to possible changes in CFR over the course of the outbreak and the possibility of an intervention affecting the CFR, we also considered a decay model where the CFR changed at the time of intervention implementation, τ_1_, with the effect of the intervention delayed by 0, 4, 7, or 10 days:

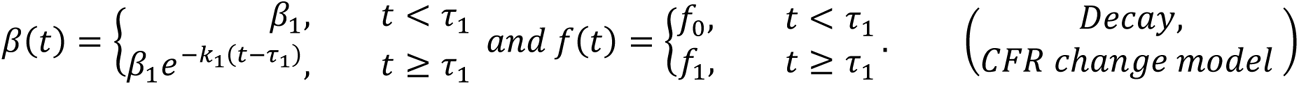

Again, MSE values were calculated for daily incidence counts and cumulative case and death counts over time to determine whether including the changing CFRs provided estimates more similar to the reported data.

#### 2. Second intervention

##### a. Decaying transmission

To examine the effect of multiple interventions, the model was parameterized with data through December 4^th^, starting with either September 20^th^ or September 24^th^, depending on which had a better fit above (September 20th – October 14th vs. September 24th – October 14th). For all scenarios in this time frame, the outbreak start date (τ_0_), the delayed effect of the first intervention (if any), and the change in CFR at first intervention (if any) were set based on the best fit model from above. The first scenario examined was the “decay model” from above, which accounted for only the first intervention, thereby acting as a baseline where the second intervention had no effect. This was parameterized with β_1_, k_1_, f_0_, and (if applicable) f_1_ estimated using MLE (Table 1). We then compared this baseline to an n = 2 version of the “independent decay model” from above, parameterizing it to include the decaying effects of both interventions on transmission rate, but not the diminished effects for the first intervention or the changing of case fatality rate (CFR) due to the second intervention (τ_1*end*_ = τ_2_, β_2_ = β_1_, *and f*_2_ = *f*_1_).

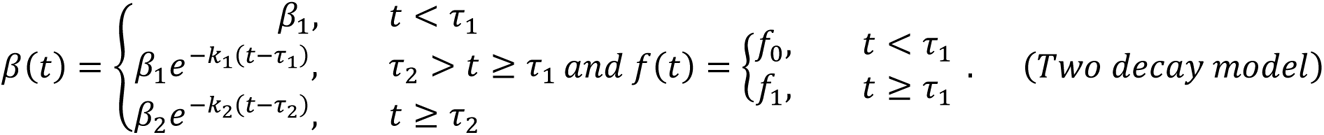

This will be referred to as the “two decay model”. MSE comparisons were made overall, as well as for the dataset up until October 15th (the dates used to examine the first intervention) and from October 15th onward (the dates used to examine the second intervention).

##### b. Delayed decay

If the “two decay model” had the better fit compared to the “decay model”, we considered parameterizations where the second intervention had its effect delayed by 4, 7, or 10 days, setting τ_2_to be October 19^th^, 22^nd^, and 25^th^, respectively (Table 1). Particularly for the implementation of a lockdown, there was the possibility of a delayed effect based on stringency, compliance rate, and enforcement.

##### c. Changing case fatality rates

We also considered a version of the “two decay model” where the CFR changed at the time the second intervention went into effect, τ_2_, with 0, 4, 7, and 10-day delayed effects:

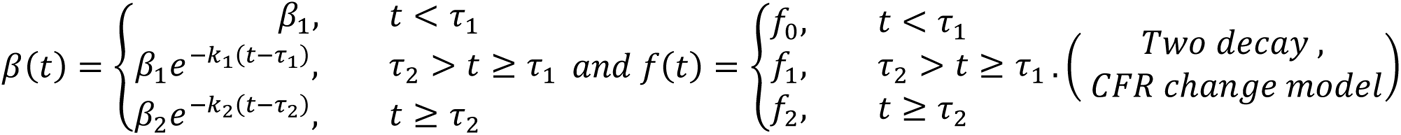

#### 3. Diminished effects

##### a. Diminished effects of the first intervention

To determine whether the effects of the first intervention diminished before the second intervention was put in place, the “independent decay model” was parameterized where the first intervention had decaying effects from time of implementation (τ_1_) up until October 15^th^ (τ_1*end*_). These decaying effects ceased and a new constant transmission rate was set from τ_1*end*_ until the date of the second intervention taking its delayed effect (τ_2_), either 4, 7, or 10 days after October 15^th^:

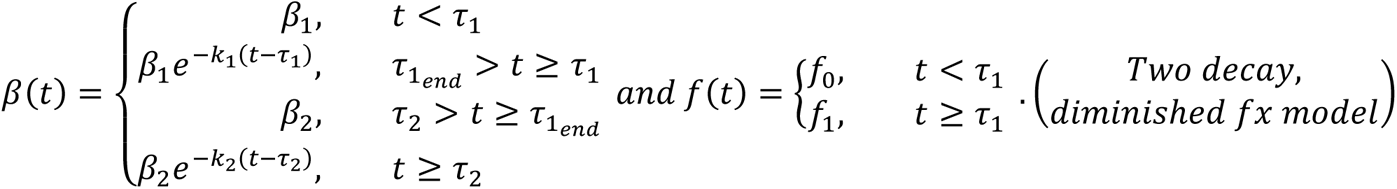

Finally, we ran a version of the n = 2 “independent decay model” that allowed for diminished effects of the first intervention, delayed effects of the second intervention, and the CFR changing at time τ_2_, when the second intervention took effect:

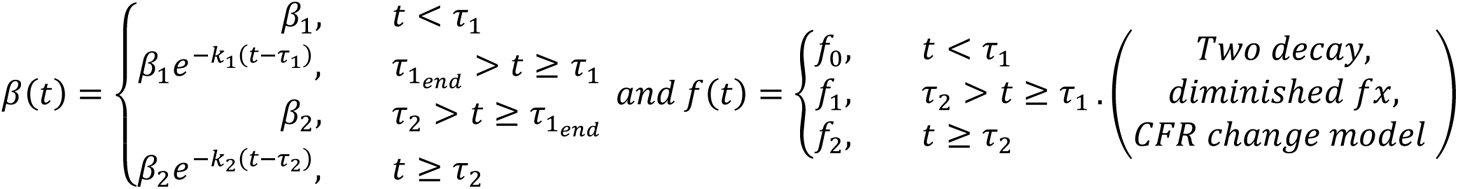

## Results

After the first case of SVD was detected on September 20, 2022, reported case counts were most similar to the MoH onset data transposed 6 days ahead (Figure 1a) (Table S1-2). However, the time between case onset and reporting started to decrease during the middle of the outbreak, with reported counts best matching onset counts transposed 4 days ahead (Table S1). By the end of the outbreak, cases were being reported within zero to two days of symptom onset (Figure 1a) (Table S2). When examining R_e_ estimates over time, the trend of the estimates from reported case data matched those estimated from the MoH onset dataset with a time lag (Figure 1b). However, the reported case count dataset produced slightly higher R_e_ estimates at the peaks and slightly lower estimates at the valleys.

**Figure 1:**
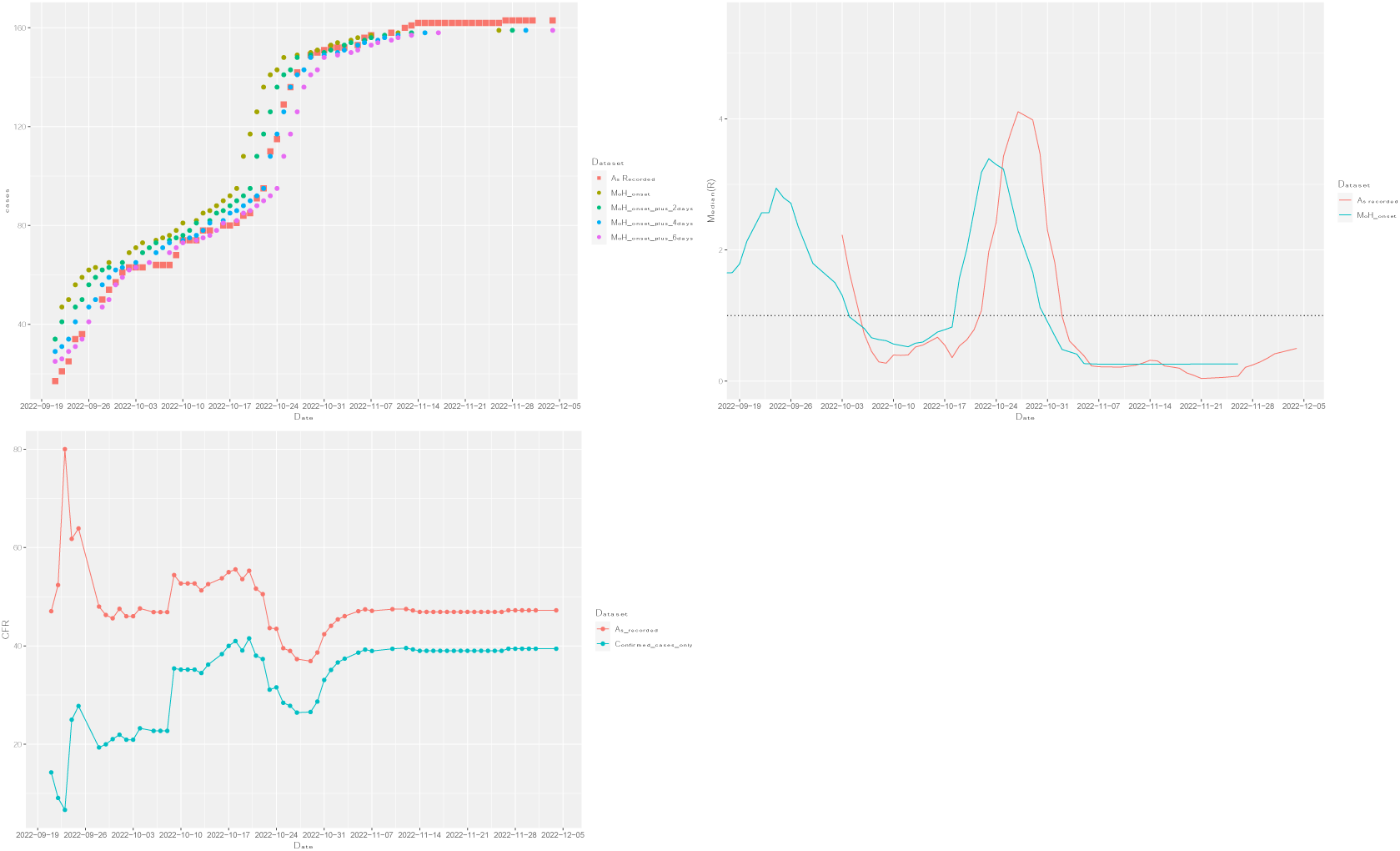
Changes over time in epidemic case counts, Re estimates, and CFR values. (a) Difference between onset and capture date over time. (b) Estimated Re over time using case counts as recorded and by onset date. (c) Case fatality ratio (CFR) values over time using as cases and only confirmed cases.

When examining CFR values, those calculated from the “confirmed reported only” dataset were lower than those calculated from the “as reported” dataset between September 22nd and October 8th (Table S3). This is likely due to a denominator inflation issue when including the probable cases from the data onboarding period (September 24th – October 14th). Indeed, by October 1st, the two sets of CFR values had similar trends, with the ratios slowly growing closer to each other as the number of cases increased and the impact of adding twenty probable cases and twenty probable deaths lessened. By the end of the outbreak, CFR was estimated at 39.43% when including only confirmed cases and 47.24% when including confirmed and probable cases — estimates that were not found to be significantly different from each other (Chi-square Test of Homogeneity: p = 0.21) (Table S3). For both sets of CFR values, estimates decreased during the periods when case counts were increasing and vice versa.

When examining the effect of the first intervention on transmission during September 20th – October 14th, the predicted values from the “decay” models (1.a.iii-iv) better fit the “as reported” dataset, as compared to the “base” models (1.a.i-ii) (Table 3). This was also the case when comparing “base” (1.b.i-ii) and “decay” (1.b.iii-iv) model fits to the dataset excluding the data onboarding period. Further, whether including or excluding the data onboarding period, the “decay” models estimated a higher transmission rate (β_1_), and therefore a higher R_0_, than the base models. While models that treated the outbreak start date (τ_0_) as a free parameter generally fit better than their fixed τ_0_counterparts (1.a.i vs. 1.a.ii, 1.a.iii vs. 1.a.iv, 1.b.i vs. 1.b.ii, 1.b.iii vs. 1.b.iv), they produced estimated start dates that were quite different from the “correct” date of August 7^th^, as reported by MoH Uganda and the CDC. The “base” models with a free τ_0_ parameter estimated earlier start dates (1.a.i: July 31st; 1.b.i: July 13th), leading to a longer underlying spread with a lower transmission rate (β_1_) and lower R_0_ estimates (Table 2). For the “decay” model starting on September 20^th^ (1.a.iii), τ_0_ was estimated to be more recent (August 28^th^) and both the transmission (β_1_=0.543) and decay rate (k_1_=0.116) were estimated to be higher than in the fixed τ_0_ model (1.a.iv: β_1_=0.396, k_1_=0.026), likely because the model was fit to the first few data points when data onboarding was occurring (Table 2) (Figure 2). Indeed, when comparing model fits with (1.a.i-iv) and without (1.b.i-iv) the data onboarding period, transmission rate (β_1_) and decay rate (k_1_) estimates were similar when the start date of the outbreak (τ_0_) was fixed at August 7th (1.a.iv: β_1_=0.396, k_1_=0.026; 1.b.iv: β_1_=0.402, k_1_=0.040) (Table 2). When the start date (τ_0_) was treated as a free parameter, however, there were large differences in parameter estimates between the two models (1.a.iii: β_1_=0.543, k_1_=0.116; 1.b.iii: β_1_=0.403, k_1_=0.040) (Table 2). Overall, the model fits with the onboarding period data (1.a.i-iv) produced predicted values more similar to the observed (“as reported”) dataset than the model fits without the onboarding period data (1.b.i-iv), both in terms of MSE and Pearson goodness-of-fit statistic values (Table 3-4). However, when examining graphs of these predicted and observed values, it seems that models 1.a.i-iv were only better at fitting the data points on September 24^th^ and 25^th^, the days immediately following the chosen data onboarding period (Figure 2). Further, the “decay” model starting on September 24^th^ with τ_0_ as a free parameter (1.b.iii) estimated the “correct” outbreak start date (August 7^th^) and had very similar estimates for β_1_and k_1_ compared to the “decay” model with fixed τ_0_ (1.b.iv) (Table 2); therefore, we used the dataset starting from September 24^th^ for all subsequent models.

**Figure 2:**
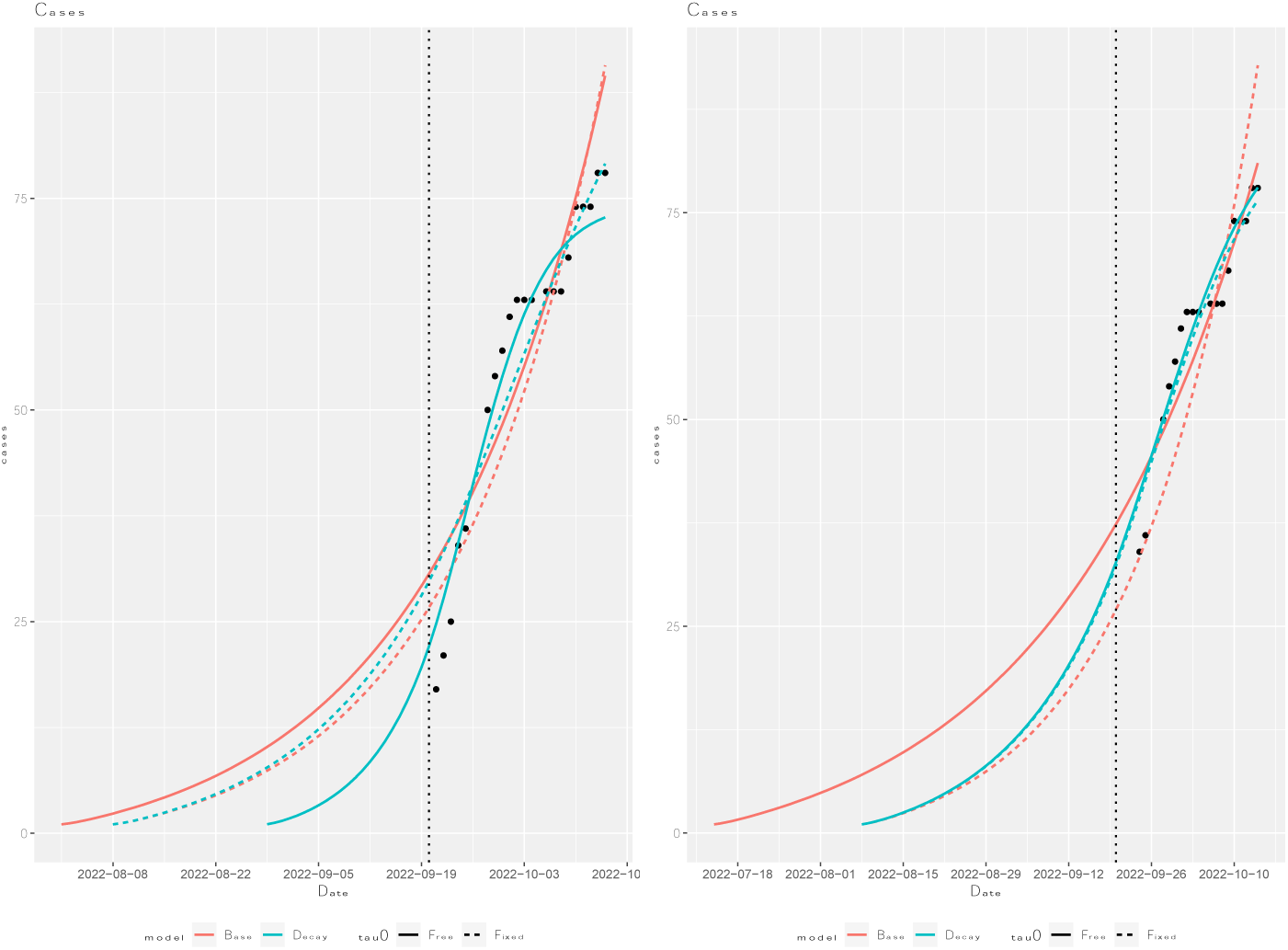
Simulated cumulative case counts for base and decay models with τ_0_as a fixed or free parameter run from (a) 9/20 – 10/14 and (b) 9/24 – 10/14.

**Table 2:** Maximum Likelihood Estimates (MLE) for each model.

| Model | Primary case date ($\tau_0$) | $\beta_1$ | $k_1$ | $\beta_2$ | $k_2$ | $f_0$ (%) | $f_1$ (%) | $f_2$ (%) | $R_0$ |
| --- | --- | --- | --- | --- | --- | --- | --- | --- | --- |
| 1.a.i | 7/31/22 | 0.374 | --- | --- | --- | 58.7 | --- | --- | 1.31 |
| 1.a.ii | 8/7/22* | 0.388 | --- | --- | --- | 60.4 | --- | --- | 1.36 |
| 1.a.iii | 8/28/22 | 0.543 | 0.116 | --- | --- | 58.7 | --- | --- | 1.90 |
| 1.a.iv | 8/7/22* | 0.396 | 0.026 | --- | --- | 57.7 | --- | --- | 1.39 |
| 1.b.i | 7/13/22 | 0.347 | --- | --- | --- | 55.8 | --- | --- | 1.21 |
| 1.b.ii | 8/7/22* | 0.389 | --- | --- | --- | 60.2 | --- | --- | 1.36 |
| 1.b.iii | 8/7/22 | 0.403 | 0.040 | --- | --- | 56.1 | --- | --- | 1.41 |
| 1.b.iv | 8/7/22* | 0.402 | 0.040 | --- | --- | 56.1 | --- | --- | 1.41 |
| 1.c.i | 8/4/22 | 0.389 | 0.057 | --- | --- | 56.0 | --- | --- | 1.36 |
| 1.c.ii | 8/7/22* | 0.400 | 0.070 | --- | --- | 56.1 | --- | --- | 1.40 |
| 1.c.iii | 7/31/22 | 0.380 | 0.081 | --- | --- | 55.9 | --- | --- | 1.33 |
| 1.c.iv | 8/7/22* | 0.399 | 0.120 | --- | --- | 55.7 | --- | --- | 1.40 |
| 1.c.v | 7/27/22 | 0.370 | 0.115 | --- | --- | 55.8 | --- | --- | 1.29 |
| 1.c.vi | 8/7/22* | 0.398 | 0.332 | --- | --- | 56.1 | --- | --- | 1.39 |
| 1.d.i | 8/22/22 | 0.479 | 0.071 | --- | --- | 84.3 | 41.5 | --- | 1.68 |
| 1.d.ii | 8/7/22* | 0.401 | 0.036 | --- | --- | 65.1 | 48.1 | --- | 1.40 |
| 1.d.iii | 8/18/22 | 0.450 | 0.108 | --- | --- | 71.8 | 41.0 | --- | 1.57 |
| 1.d.iv | 8/7/22* | 0.399 | 0.061 | --- | --- | 63.1 | 46.6 | --- | 1.40 |
| 1.d.v | 8/10/22 | 0.408 | 0.126 | --- | --- | 63.4 | 42.8 | --- | 1.43 |
| 1.d.vi | 8/7/22* | 0.398 | 0.106 | --- | --- | 61.9 | 44.6 | --- | 1.39 |
| 1.d.vii | 8/1/22 | 0.381 | 0.154 | --- | --- | 58.8 | 46.0 | --- | 1.33 |
| 1.d.viii | 8/7/22* | 0.397 | 0.246 | --- | --- | 60.6 | 42.1 | --- | 1.39 |
| 2.a.i | 8/7/22* | 0.392 | 0.018 | --- | --- | 50.0 | --- | --- | 1.37 |
| 2.a.ii | 8/7/22* | 0.395 | 0.032 | --- | 0.037 | 49.9 | --- | --- | 1.38 |
| 2.b.i | 8/7/22* | 0.393 | 0.023 | --- | 0.051 | 49.9 | --- | --- | 1.38 |
| 2.b.ii | 8/7/22* | 0.392 | 0.020 | --- | 0.068 | 49.9 | --- | --- | 1.37 |
| 2.b.iii | 8/7/22* | 0.391 | 0.017 | --- | 0.097 | 49.9 | --- | --- | 1.37 |
| 2.c.i | 8/7/22* | 0.392 | 0.025 | --- | 0.037 | 56.3 | --- | 39.4 | 1.37 |
| 2.c.ii | 8/7/22* | 0.391 | 0.019 | --- | 0.051 | 55.2 | --- | 37.9 | 1.37 |
| 2.c.iii | 8/7/22* | 0.390 | 0.017 | --- | 0.067 | 54.5 | --- | 36.6 | 1.37 |
| 2.c.iv | 8/7/22* | 0.390 | 0.015 | --- | 0.093 | 53.8 | --- | 35.1 | 1.36 |
| 3.a.i | 8/7/22* | 0.402 | 0.127 | 1.252 | 0.257 | 49.6 | --- | --- | 1.41 |
| 3.a.ii | 8/7/22* | 0.401 | 0.116 | 0.994 | 0.419 | 49.6 | --- | --- | 1.40 |
| 3.a.iii | 8/7/22* | 0.401 | 0.108 | 0.883 | 4.150 | 49.7 | --- | --- | 1.40 |
| 3.b.i | 8/7/22* | 0.397 | 0.091 | 1.034 | 0.212 | 56.7 | --- | 23.7 | 1.39 |
| 3.b.ii | 8/7/22* | 0.399 | 0.111 | 1.026 | 0.428 | 54.3 | --- | 36.8 | 1.40 |
| 3.b.iii | 8/7/22* | 0.399 | 0.106 | 0.919 | 5.303 | 54.9 | --- | 35.2 | 1.40 |

**Table 3:** Mean Squared Error (MSE) values for cumulative cases, cumulative deaths, and daily incidence during the periods from 9/24 – 10/14, 10/15 – 12/4, and overall (9/24-12/4). Lowest MSE value is in red for each section of models.

| Model | Overall MSE (9/24 – 12/4) |  |  | 9/24* – 10/14 MSE |  |  | 10/15 – 12/4 MSE |  |  |
| --- | --- | --- | --- | --- | --- | --- | --- | --- | --- |
|  | Cases | Deaths | Incidence | Cases | Deaths | Incidence | Cases | Deaths | Incidence |
| 1.a.i | --- | --- | --- | 44.96 | 7.1 | 15.86 | --- | --- | --- |
| 1.a.ii | --- | --- | --- | 70.12 | 11.97 | 16.5 | --- | --- | --- |
| 1.a.iii | --- | --- | --- | 10.76 | 11.96 | 12.18 | --- | --- | --- |
| 1.a.iv | --- | --- | --- | 20.35 | 5.12 | 14.1 | --- | --- | --- |
| 1.b.i | --- | --- | --- | 21.1 | 2.78 | 15.6 | --- | --- | --- |
| 1.b.ii | --- | --- | --- | 72.0 | 12.9 | 16.6 | --- | --- | --- |
| 1.b.iii | --- | --- | --- | 13.1 | 4.62 | 13.8† | --- | --- | --- |
| 1.b.iv | --- | --- | --- | 12.5† | 4.15 | 14.0 | --- | --- | --- |
| 1.c.i | --- | --- | --- | 17.1 | 5.00 | 14.1 | --- | --- | --- |
| 1.c.ii | --- | --- | --- | 11.8† | 4.79 | 13.7 | --- | --- | --- |
| 1.c.iii | --- | --- | --- | 13.3 | 4.35 | 14 | --- | --- | --- |
| 1.c.iv | --- | --- | --- | 11.8† | 5.53 | 13.5† | --- | --- | --- |
| 1.c.v | --- | --- | --- | 14.9 | 3.82† | 14.6 | --- | --- | --- |
| 1.c.vi | --- | --- | --- | 14.3 | 6.41 | 13.9 | --- | --- | --- |
| 1.d.i | --- | --- | --- | 13.8 | 5.81 | 12.9 | --- | --- | --- |
| 1.d.ii | --- | --- | --- | 12.7 | 3.62† | 13.9 | --- | --- | --- |
| 1.d.iii | --- | --- | --- | 12.6 | 5.42 | 12.5† | --- | --- | --- |
| 1.d.iv | --- | --- | --- | 12† | 3.94 | 13.7 | --- | --- | --- |
| 1.d.v | --- | --- | --- | 12† | 4.84 | 13.4 | --- | --- | --- |
| 1.d.vi | --- | --- | --- | 12.1 | 4.4 | 13.6 | --- | --- | --- |
| 1.d.vii | --- | --- | --- | 14.4 | 4.2 | 14.3 | --- | --- | --- |
| 1.d.viii | --- | --- | --- | 14.4 | 5.18 | 14.0 | --- | --- | --- |
| 2.a.i | 101.3 | 13.41 | 10.97 | 46.8 | 25.46 | 14.95 | 124.1 | 8.36 | 9.31 |
| 2.a.ii | 74.5 | 12.6 | 10.26 | 42 | 17.6 | 14.25 | 88 | 10.4 | 8.59 |
| 2.b.i | 75.3† | 12.1 | 10.29† | 43.3† | 23† | 14.67† | 88.7† | 7.6 | 8.46 |
| 2.b.ii | 87.2 | 11.31 | 10.59 | 43.6 | 26.34 | 14.83 | 105.5 | 5.02 | 8.82 |
| 2.b.iii | 81.7 | 12.7 | 10.83 | 45.5 | 29.44 | 15 | 96.9 | 5.49 | 9.08 |
| 2.c.i | 67.1 | 8.98 | 10.21 | 37.5† | 9.29 | 14.58† | 79.4 | 8.85† | 8.38 |
| 2.c.ii | 65.7 | 11 | 10.15 | 43.2 | 12.2 | 14.88 | 75.2 | 10.5 | 8.18 |
| 2.c.iii | 73.2 | 12.4 | 10.43 | 45.9 | 15.9 | 14.98 | 84.6 | 10.9 | 8.52 |
| 2.c.iv | 68.4 | 14.4 | 10.64 | 47.8 | 17.7 | 15.11 | 77 | 13.1 | 8.77 |
| 3.a.i | 15.3 | 10.52 | 7.31 | 23 | 9.52 | 13.21 | 12.1 | 10.94 | 4.84 |
| 3.a.ii | 19.8 | 9.17 | 6.8 | 19.3 | 10.95 | 13.29 | 20 | 8.42 | 4.08 |
| 3.a.iii | 17 | 10.6 | 6.48 | 22.5 | 10.1 | 13.26 | 14.6 | 10.8 | 3.64 |
| 3.b.i | 13.4 | 4.12 | 7.59 | 13.1 | 5.64 | 13.65 | 13.6 | 3.49 | 5.05 |
| 3.b.ii | 10.21 | 4.89 | 6.65 | 12.19 | 5.62 | 13.5 | 9.39 | 4.58 | 3.78 |
| 3.b.iii | 11.6 | 6.59 | 6.4 | 12.8 | 5.65 | 13.48† | 11.1 | 6.99 | 3.44 |

**Table 4:** Goodness of fit for case counts during the periods from 9/24 – 10/14, 10/15 – 12/4, and overall (9/24-12/4). Pearson goodness-of-fit statistic, df, and p value are provided for each model. Lowest chi-squared statistic (and highest p-value) is in red for each section of models.

| Model | Overall MSE (9/24 – 12/4) |  |  | 9/24* – 10/14 MSE |  |  | 10/15 – 12/4 MSE |  |  |
| --- | --- | --- | --- | --- | --- | --- | --- | --- | --- |
|  | X<sup>2</sup> | df | p-value | X<sup>2</sup> | df | p-value | X<sup>2</sup> | df | p-value |
| 1.a.i | --- | --- | --- | 13.66 | 17 | 0.6908 | --- | --- | --- |
| 1.a.ii | --- | --- | --- | 23.73 | 17 | 0.1269 | --- | --- | --- |
| 1.a.iii | --- | --- | --- | 3.01 | 17 | 0.9999 | --- | --- | --- |
| 1.a.iv | --- | --- | --- | 7.04 | 17 | 0.9830 | --- | --- | --- |
| 1.b.i | --- | --- | --- | 7.48 | 17 | 0.9764 | --- | --- | --- |
| 1.b.ii | --- | --- | --- | 22.47 | 17 | 0.1672 | --- | --- | --- |
| 1.b.iii | --- | --- | --- | 4.50 | 17 | 0.9989 | --- | --- | --- |
| 1.b.iv | --- | --- | --- | 4.34† | 17 | 0.9991† | --- | --- | --- |
| 1.c.i | --- | --- | --- | 5.62 | 17 | 0.9954 | --- | --- | --- |
| 1.c.ii | --- | --- | --- | 3.94 | 17 | 0.9995 | --- | --- | --- |
| 1.c.iii | --- | --- | --- | 4.51 | 17 | 0.9988 | --- | --- | --- |
| 1.c.iv | --- | --- | --- | 3.76† | 17 | 0.9997† | --- | --- | --- |
| 1.c.v | --- | --- | --- | 5.11 | 17 | 0.9974 | --- | --- | --- |
| 1.c.vi | --- | --- | --- | 4.35 | 17 | 0.9991 | --- | --- | --- |
| 1.d.i | --- | --- | --- | 4.28 | 17 | 0.9992 | --- | --- | --- |
| 1.d.ii | --- | --- | --- | 4.39 | 17 | 0.9990 | --- | --- | --- |
| 1.d.iii | --- | --- | --- | 3.51† | 17 | 0.9998† | --- | --- | --- |
| 1.d.iv | --- | --- | --- | 4.02 | 17 | 0.9995 | --- | --- | --- |
| 1.d.v | --- | --- | --- | 3.74 | 17 | 0.9997 | --- | --- | --- |
| 1.d.vi | --- | --- | --- | 3.89 | 17 | 0.9996 | --- | --- | --- |
| 1.d.vii | --- | --- | --- | 4.72 | 17 | 0.9985 | --- | --- | --- |
| 1.d.viii | --- | --- | --- | 4.46 | 17 | 0.9989 | --- | --- | --- |
| 2.a.i | 58.82 | 60 | 0.5189 | 14.03 | 17 | 0.6647 | 44.79 | 42 | 0.3557 |
| 2.a.ii | 44.06 | 60 | 0.9390 | 11.15 | 17 | 0.8486 | 32.91 | 42 | 0.8414 |
| 2.b.i | 45.77† | 60 | 0.9126† | 12.53† | 17 | 0.7673† | 33.24† | 42 | 0.8309 |
| 2.b.ii | 51.72 | 60 | 0.7681 | 13.40 | 17 | 0.7090 | 38.32 | 42 | 0.6335 |
| 2.b.iii | 50.55 | 60 | 0.8027 | 14.76 | 17 | 0.6124 | 35.78 | 42 | 0.7394 |
| 2.c.i | 41.52 | 60 | 0.9669 | 12.13 | 17 | 0.7920 | 29.38 | 42 | 0.9292 |
| 2.c.ii | 42.68 | 60 | 0.9558 | 14.28 | 17 | 0.6474 | 28.40 | 42 | 0.9461 |
| 2.c.iii | 47.37 | 60 | 0.8818 | 16.02 | 17 | 0.5224 | 31.35 | 42 | 0.8857 |
| 2.c.iv | 45.91 | 60 | 0.9102 | 16.40 | 17 | 0.4959 | 29.51 | 42 | 0.9269 |
| 3.a.i | 10.83 | 60 | 1.0000 | 6.66 | 17 | 0.9876 | 4.17 | 42 | 1.0000 |
| 3.a.ii | 11.71 | 60 | 1.0000 | 5.64 | 17 | 0.9953 | 6.07 | 42 | 1.0000 |
| 3.a.iii | 11.04 | 60 | 1.0000 | 6.38 | 17 | 0.9903 | 4.66 | 42 | 1.0000 |
| 3.b.i | 8.92 | 60 | 1.0000 | 4.28 | 17 | 1.0000 | 4.64 | 42 | 1.0000 |
| 3.b.ii | 6.79 | 60 | 1.0000 | 3.85 | 17 | 1.0000 | 2.94 | 42 | 1.0000 |
| 3.b.iii | 7.49 | 60 | 1.0000 | 4.00 | 17 | 1.0000 | 3.50 | 42 | 1.0000 |

When delaying the effect of the first intervention by 4, 7, or 10 days with the outbreak start date (τ_0_) set at August 7^th^ (1.c.ii, 1.c.iv, 1.c.vi), transmission rate (β_1_) estimates were very similar (1.c.ii: 0.400; 1.c.iv: 0.399; 1.c.vi:0.398). However, the longer the delay of effect, the higher the decay rate (k_1_) estimates were, with the 4-day delay model (1.c.ii) yielding a decay rate estimate of 0.070 and the 10-day decay model (1.c.vi) yielding a decay rate estimate of 0.332 (Figure 3) (Table 2). When the outbreak start date (τ_0_) was treated as a free parameter, the 4-(1.c.i), 7-(1.c.iii), and 10-day (1.c.v) “delay” models estimated the date to be 3, 7, and 11 days earlier than August 7th, respectively (Table 2). As these earlier first case dates provided an increased time for the disease to spread before intervention, transmission (β_1_) estimates were lowered (1.c.i: 0.389; 1.c.iii: 0.380; 1.c.v:0.370), resulting in lower R_0_ estimates (1.c.i: 1.36; 1.c.iii: 1.33; 1.c.v:1.29) (Figure 2) (Table 2).

**Figure 3:**
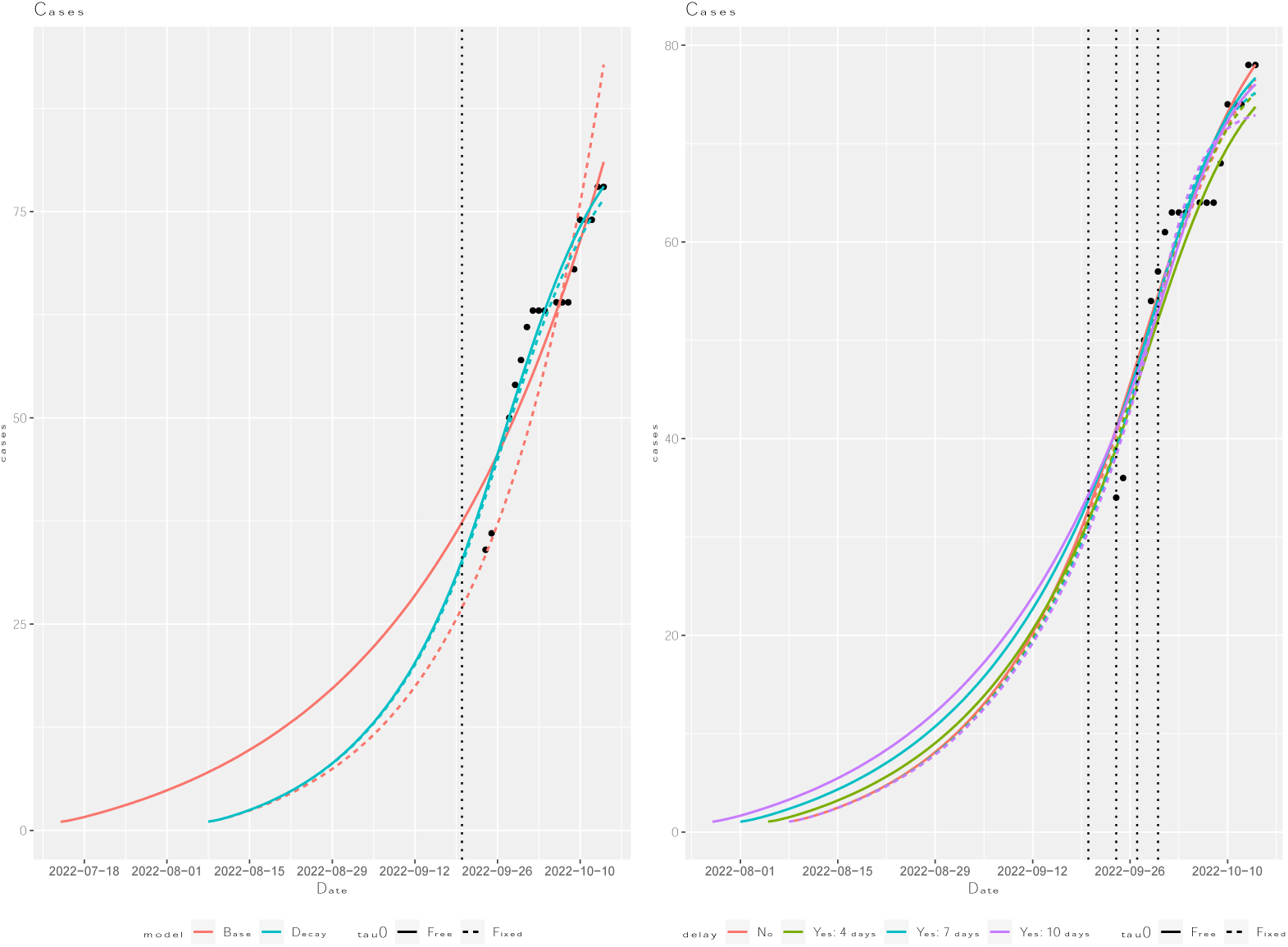
Simulated cumulative case counts for models run from 9/24 – 10/14 (a) base vs. decay and (b) decay with various delay amounts.

Allowing for the CFR (f) to change when the first intervention took effect (1.d.i-vi) had little impact on the fit of the models (Table 3-4). Interestingly, when the outbreak start date was treated as a free parameter for the “decay, CFR change” models (1.d.i, 1.d.iii, 1.d.v, 1.d.vii), the estimated start date of the outbreak was later than August 7th, the opposite effect than seen with “decay” models with delay and a constant CFR (1.d.ii, 1.d.iv, 1.d.vi, 1.d.viii) (Table 2).

However, similarly to the models with one CFR, increased delay of intervention effects moved the estimated start date earlier for the “CFR change” models. That is, the 0-day “delay” model (1.d.i) estimated the start date as August 22nd, with increased delay moving the start date earlier (1.d.iii: August 18th; 1.d.v: August 10th; 1.d.vii: August 1st). Increased delay of intervention effects and earlier estimated outbreak start dates had corresponding increases in decay rate (k_1_) estimates (1.d.i: 0.071; 1.d.iii: 0.108; 1.d.v: 0.126; 1.d.vii: 0.154) and decreases in transmission rate (β_1_) estimates (1.d.i: 0.479; 1.d.iii: 0.450; 1.d.v: 0.408; 1.d.vii: 0.381), as seen for the models with one CFR (Table 2). Overall, the 4-and 7-day delayed “decay” models with a constant CFR and fixed outbreak start date (τ_0_) (1.d.iv and 1.d.vi) yielded predicted cumulative case values that best fit the observed data, with the 7-day delayed version (1.d.vi) yielding a better fit for deaths (Table 3). Therefore, in all subsequent models, we assumed the first intervention took effect 7 days after its implementation and there was no change in case fatality rate (*f*_1_ = *f*_0_).

When comparing the model where both interventions have decaying effects on transmission (2.a.ii) to that where only the first intervention has an effect (2.a.i), we found that the former produced a significantly better fit to the “as reported” dataset — not just overall and for the second time period (October 15th – December 4th), but also for the first time period (September 24th – October 14th) (Table 3). This “two decay” model (2.a.ii) estimated a slightly lower transmission rate (β_1_= 0.395) and a smaller decay rate for the first intervention (k_1_ = 0.032) compared to the “decay” model for September 24th – October 14th (1.d.iv: β_1_ = 0.399; k_1_= 0.061) (Table 2). Accounting for a delayed effect of the second intervention with no change in CFR (2.b.i-iii) did not provide a better fit; however, the model with a 4-day delay and changing CFR (2.c.ii) did fit the data better (Figure 4) (Table 3). For the 0, 4, 7, and 10-day delayed “two decay, CFR change” models (2.c.i-iv), the estimates of β_1_ and k_1_ were slightly lower than their “two decay” model counterparts (2.a.ii, 2.b.i-iii), but the estimates for k_2_ were comparable (Table 2). For all the “two decay, CFR change” models (2.c.i-iv), the CFR started high (53.6 – 56.3%) then decreased (to 35.1-39.4%) (Table 2). For all the “two decay” models (2.a.ii, 2.b.i-iii, 2.c.i-iv), regardless of days delayed or number of case fatality rates, the R_0_ value stayed in the window of 1.36 to 1.38 (Table 2).

**Figure 4:**
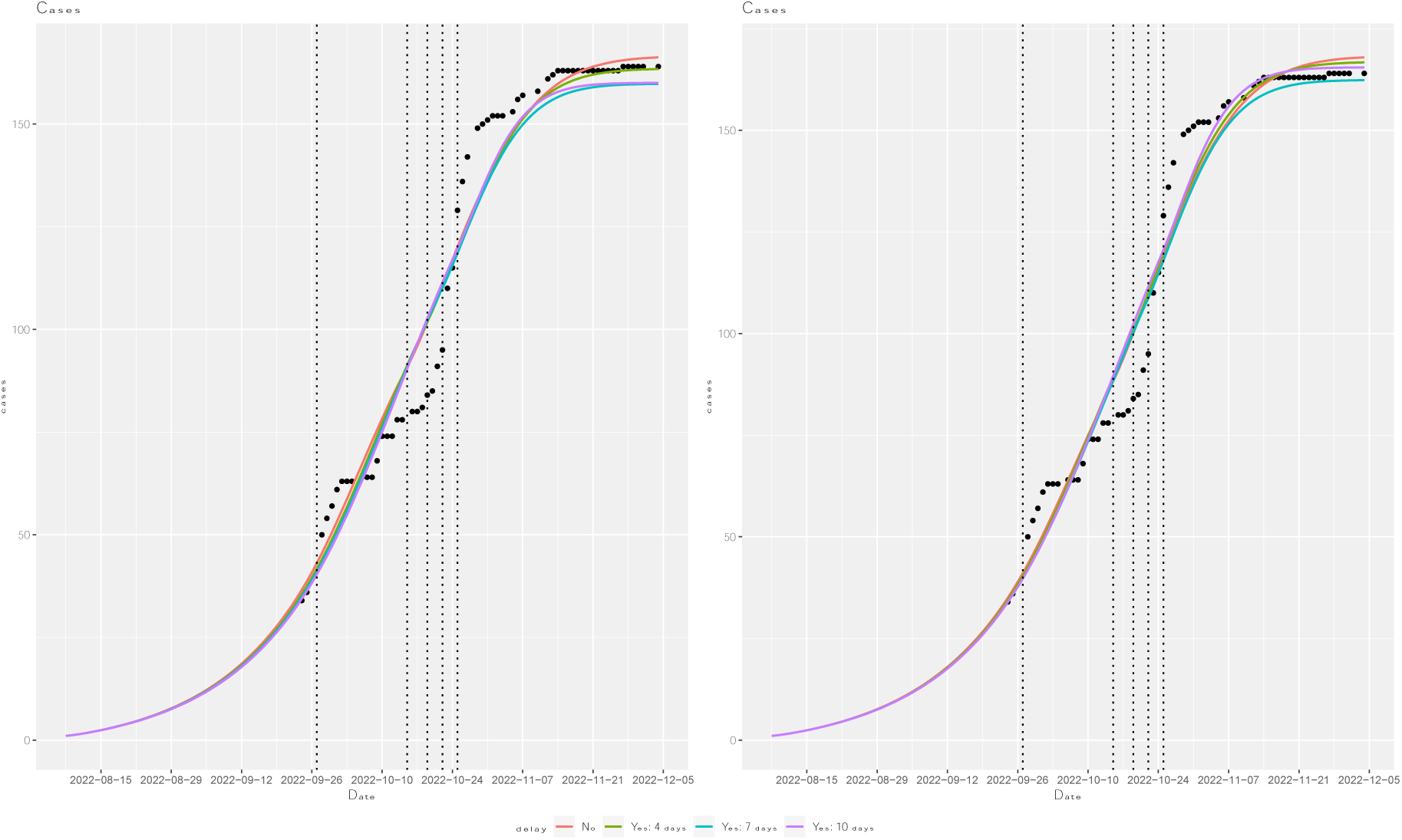
Simulated cumulative case counts for ‘Two decay’ models run from 9/24 – 12/4 with various delay amounts and (a) one or (b) two fatality rates.

While the “two decay” models provided good fits overall, they were unable to capture the trend seen in the data around October 15^th^ – October 25^th^, where the rate of new cases decreased and then rapidly increased (Figure 4). This trend was captured, however, when accounting for a diminished effect of the first intervention on transmission (Figure 5). Indeed, the “two decay, diminished fx” models (3.a.i-iii) all produced considerably lower MSE values for cumulative cases and deaths, overall and by time period. Particularly for the October 15th – December 4th time period, MSE values for cumulative cases dropped from 75.2-84.6 (2.c.ii-iv) to 12.1-20 (3.a.i-iii) and MSE values for deaths were cut in half, going from 8.18-8.77 (2.c.ii-iv) to 3.64-4.84 (3.a.i-iii) (Table 3). All three of these “two decay, diminished fx” models (3.a.i-ii) yielded slightly higher initial transmission estimates (β_1_) than their “two decay” counterparts (2.c.ii-iv); however, they also yielded increased decay rates (k_1_ and k_2_) (Table 2). None of these models produced the best fit for incidence, case, and death counts; rather, each one had the best MSE value for either daily incidence counts, cumulative case counts, or cumulative death counts (Table 3). This tradeoff between fitting the model to cumulative cases or deaths was also apparent in the graph, where the model with the best fit for cumulative case counts had the worst fit for cumulative death counts, particularly at the end of the outbreak (Figure 5) (Figure S2). The “two decay, diminished fx, CFR change” models (3.b.i-iii) all produced better fits for cumulative cases and deaths than the “two decay, diminished fx” models (3.a.i-iii). Particularly, the “CFR change” models yielded better fits for the death counts during October 10^th^ – October 31^st^ (Figure 5). There was, however, no clear best-fit between the “CFR change” models with 4, 7, and 10-day delayed decays (Table 3). For all the “two decay, diminished fx” models, regardless of days delayed or number of CFRs (3.a.i-iii, 3.b.i-iii), the R_0_ value stayed in the window of 1.39 to 1.41 (Table 2). While this was slightly above the values of the “two decay” models that did not account for diminished effects of the first intervention (2.a.ii, 2.b.i-iii, 2.c.i-iv), it was the same as the R_0_ estimate from all the “decay” models run on the first portion of the data (September 24th – October 14th) where outbreak start date was set or estimated as August 7th (1.b.iii-iv, 1.c.ii, 1.c.iv, 1.c.vi, 1.d.ii, 1.d.iv, 1.d.vi, 1.d.viii) (Table 2).

**Figure 5:**
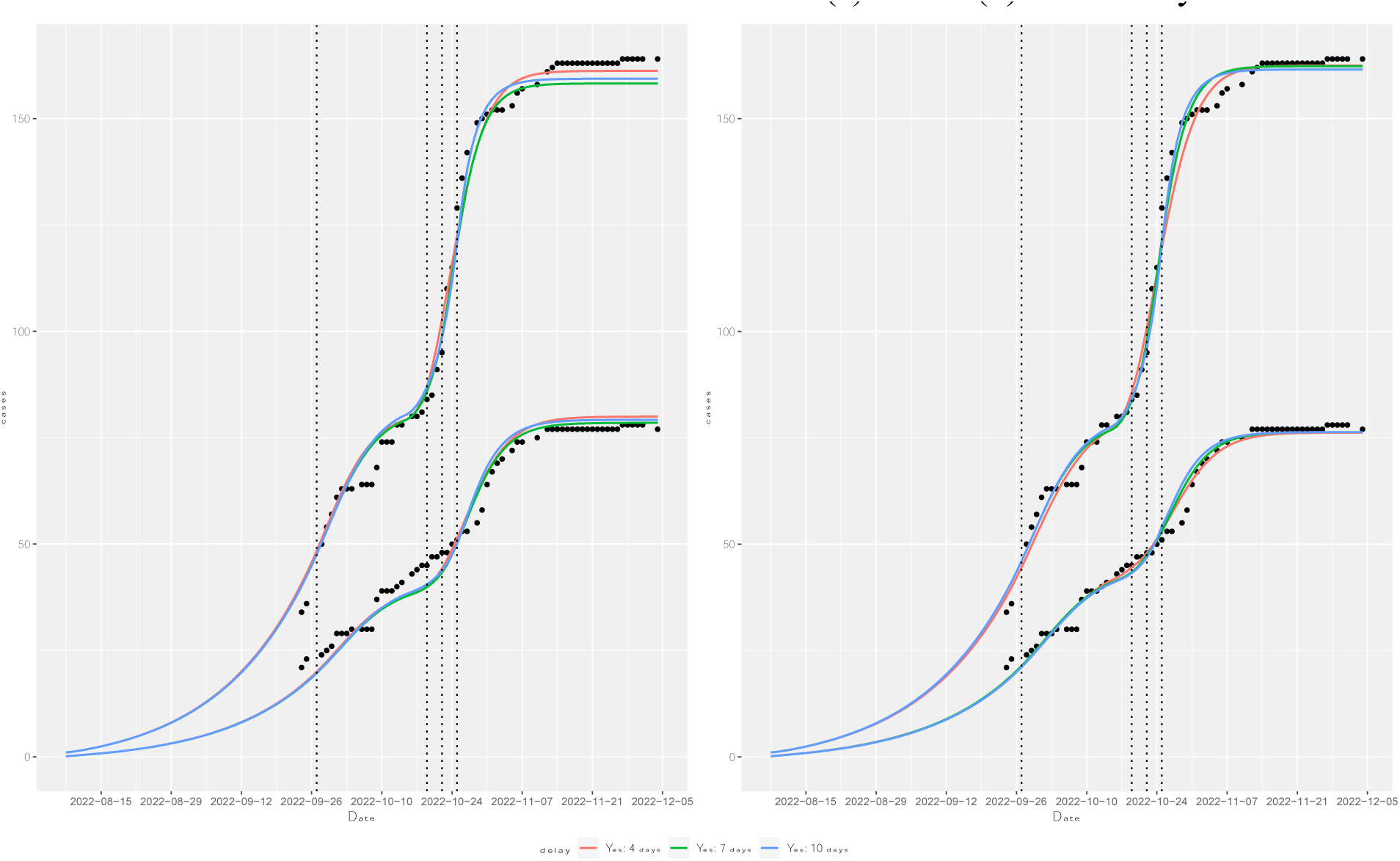
Simulated cumulative case and death counts for ‘Two decay’ models run from 9/24 – 12/4 with diminished effects of the first intervention and (a) one or (b) two fatality rates.

## Discussion

Modeling diseases throughout the course of an outbreak is an important step to understand how quickly the disease is spreading, whether interventions are having an effect, and if additional or alternative interventions should be put into place. Unfortunately, there are many challenges in such modeling endeavors, particularly during the early phase of the outbreak. Here, we present a modeling framework that can account for delayed or diminished effects of multiple interventions, changing case fatality rates, and unknown outbreak start dates. We then fit this modeling framework to data from the 2022 SVD outbreak in Uganda to examine how accounting, or not accounting, for these factors can affect estimates of disease spread.

Our work suggests that treating the outbreak start date (the date of the primary case’s symptom onset) as a free parameter — as is often necessary early in an outbreak when this date is unknown — can have a significant effect on the estimates of disease transmission. Further, the effects of treating outbreak start date as a free versus a fixed parameter vary depending on model type and data used. When the data onboarding period was included, the “base” models with known and unknown outbreak start dates had similar parameter estimates, whereas the “decay” models had quite different estimates. Comparatively, when the onboarding data period was excluded, the “decay” models had similar parameter estimates and the outbreak start date was accurately estimated when treated as a free parameter. This lack of uniform effect can make it difficult to account for the possible effects of having an unidentified primary case, and therefore an unknown outbreak start date.

Whether or not onboarding data are included, the decay model with a fixed outbreak start date estimated a higher transmission rate than the base model; this suggests that excluding intervention effects in early phase models may produce underestimated values for R_0_. The amount of delay included in a model before the effects of an intervention are seen can also impact the R_0_ estimate. R0 estimates were similar for delay models with a fixed outbreak start date; however, when the start date was unknown, models with longer delays estimated an earlier, more gradual beginning of the outbreak. For both fixed and free outbreak start dates, the longer the delay in observed effect after the implementation of an intervention, the more pronounced that decaying effect can be on transmission. Including the diminished effect of an intervention can also yield more pronounced decaying effects on transmission, but initial transmission rate estimates are higher. Not accounting for these diminished effects when modeling an outbreak could cause underestimates of both the intensity of disease spread and the efficacy of an intervention.

Allowing for a change in fatality rate later in an outbreak can also provide a better model fit. For diseases with a high CFR, such as SVD, early CFR estimates are likely to be inflated from retrospective case/death reporting, with an eventual decrease over the course of the outbreak (12, 13). This can result in CFR being underestimated during increases in case counts due to the previously reported underestimation bias during outbreak peaks from the delay between disease onset and outcome (i.e., death or recovery) (12, 13, 62). Indeed, there can be equivalent time-based changes for diseases with lower CFRs as well, such as the complete decoupling in trends between reported cases and deaths due to underreporting of the former during the COVID-19 outbreak (63, 64). CFR values can also decrease during the outbreak due to the impact of interventions, such as contact tracing efforts. These efforts can decrease the time between symptom onset and case reporting, increasing an individual’s chance of recovery and causing a change in the relationship between case counts and death counts due to quicker case detection. This change in relationship between case and death counts highlights the difficulty of using “date of reporting” data rather than “date of onset” data as well as the difficulty in trying to adjust the former to better reflect the latter.

Changes in case and death count trends over the course of the outbreak can also affect the maximum likelihood estimates (MLE) for models, as case and death counts contribute equally to the estimate. This tradeoff between fitting to cases or deaths can especially be seen with our diminished effects models, where a different model’s predictions were closest to the observed counts for daily incidence and cumulative cases and deaths. Accounting for the outbreak start date, changing case fatality rates, and diminished intervention effects can have important impacts on the transmission and decay rate estimates, particularly at the beginning of an outbreak. While excluding these factors can lead to large differences in R_0_ estimates, their inclusion makes it possible to accurately estimate R_0_ at the beginning of an outbreak, as seen for our best-fit decay model with a 7-day delay run on the short dataset (September 24^th^ – October 14^th^). Interestingly, all models run on the full dataset (September 24^th^ – December 4^th^) had precise R_0_ estimates, demonstrating the robustness of R_0_ as a measure of disease spread when examining data from the entire outbreak.

As is the case with any population-level disease model, simplifying assumptions had to be made for our models, and thus, our R_0_ estimates must be considered carefully. However, given that models like these are often the ones used at the beginning of an outbreak to provide early R_0_ estimates and policy recommendations, we believe it is important to provide the best possible framework for producing accurate estimates though an SEIDR model. Within the setting of our case study, these estimates should not be used as definite values for this SVD outbreak as incubation and infectiousness rate were fixed based on sparse empirical data. We chose to use these values to minimize the already large number of free parameters in our model, particularly given that our primary goal was to compare models and develop a modeling framework rather than to provide exact estimates of R_0_ for the outbreak. Finally, as with any dataset being reported and compiled at the beginning of an outbreak, there were trivial inconsistencies in the data reported, as mentioned in the Methods section and further cataloged in Text S1.

Future steps for producing accurate and robust estimations of epidemic potential include examining more case studies to determine whether the comparative effects seen here are consistent across outbreaks. When examining future case studies, alternative methods for calculating MLE should be considered, such as using only cumulative cases, a weighted value where cumulative cases are given more importance than cumulative deaths, or a value that accounts for the eventual lagging trend of death counts. Determining best-fit models using MSE by time period was also an imperfect method. For example, when comparing the decay models for the datasets with and without data from the onboarding period, the first had a better MSE value, but this turns out to be only due to the first two data points, with the second version fitting better for the rest of the data points. It may be worth considering daily MSE values or taking graphical outputs into account rather than relying on MSE when determining the best-fit model.

Modeling the beginning of an outbreak is a difficult endeavor — accounting for sparse data, retrospective case reporting, unknown primary case date, and more. Regardless, it is necessary to have tools that can estimate transmission intensity and determine the efficacy of multiple intervention efforts. Here, we provide a modeling framework that aims to account for these issues and we use a case study of SVD in Uganda to determine how inclusion of these effects can impact R_0_ estimates. While many challenges remain in modeling disease spread throughout the course of an outbreak, this work provides an important step on the path to accurate and robust estimations of epidemic potential.

## Data Availability

The datasets analyzed during this study are all publicly available and references throughout the manuscript.

## Supplementary Materials

Text S1: Data inconsistency issues

– Situation Report 20 (10/9/22):
  – big jump in CFR following data cleaning and harmonization, 6 outcomes have been updated as deaths’
– Situation Report 68 (12/4/22):
  – “following a data reconciliation exercise between case management and surveillance, one case previously classified as ‘dead’ is re-classified as ‘recovered’”
– Situation Report 93 (1/11/23):
  – graph states that 20 probable deaths occurred before Sept 13^th^, but only 18 cases seen
– Contact tracing values changing after contacts have ceased being added:
  – Situation Report 77 (12/18/22):
    – top lists 4525 contacts completing 21-day follow-up at top, tables has 4793 contacts listed and 4524 contacts listed as completing follow-up
  – Situation Report 78 (12/19/22):
    – top lists 4525 contacts completing 21-day follow-up at top, has all contacts listed as completing follow-up in table (totals 4793)
  – Situation Report 80 (12/23/22):
    – now listed 4793 contacts completing 21-day follow-up at top, has all contacts listed as completing follow-up in table
  – Situation Report 71 (12/11/22):
    – in contact tracing table, Jinja has 536 contacts listed and 392 contacts that completed 21-day follow-up
  – Situation Report 73 (12/13/22):
    – in contact tracing table, Jinja has 536 contacts listed and all contacts are now listed as completing 21-day follow-up (note that there are currently only 6 contacts listed as active, so unlikely all contacts in Jinja finished follow-up between these two reports)

→ When calculating the total contacts listed and having finished 21-day follow-up, 4793 used for total listed, 4380 listed as finishing follow-up (4524 listed in Situation Report 77 table – 144 added to Jinja in Situation Report 73 table) → 91% completed follow-up

Text S2: Data sources

– Contact tracing intervention data is from the situation reports https://www.afro.who.int/countries/publications?country=879
– Instating of lockdown is from presidential address https://www.yowerikmuseveni.com/address-nation-measures-stem-spread-ebola
– Extension of lockdown is from another presidential address https://www.mediacentre.go.ug/media/he-presidents-speech-ebola-virus-disease-outbreak
– Case and death count data:
  – Sept 19 → WHO Africa news report – outbreak declaration https://www.afro.who.int/countries/uganda/news/uganda-declares-ebola-virus-disease-outbreak
  – Sept 21 → (WHO Africa news report) https://www.afro.who.int/countries/uganda/news/who-bolsters-ebola-disease-outbreak-response-uganda
  – Sept 22-25 → Ministry of Health Uganda twitter updates https://twitter.com/MinofHealthUG
  – Sept 27 → WHO Africa twitter update https://twitter.com/WHOAFRO
  – Sept 29 → Situation report #10
  – Sept 30 → Ministry of Health Uganda twitter update (posted Oct 2nd but for cases since Sept 20th) https://twitter.com/MinofHealthUG
  – Oct 1 onward → situation reports 12 and up https://www.afro.who.int/countries/publications?country=879 (landing page)

**Figure S1:**
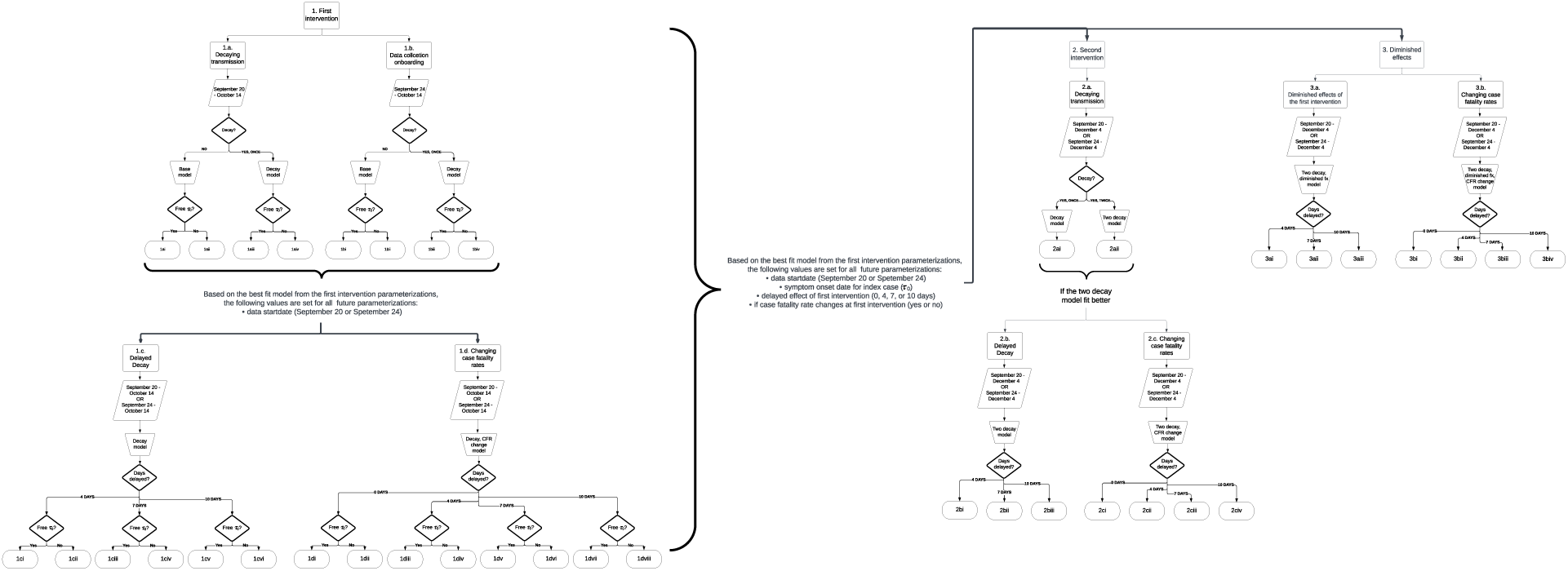
Flowchart depicting the model parameterization process.

**Figure S2:**
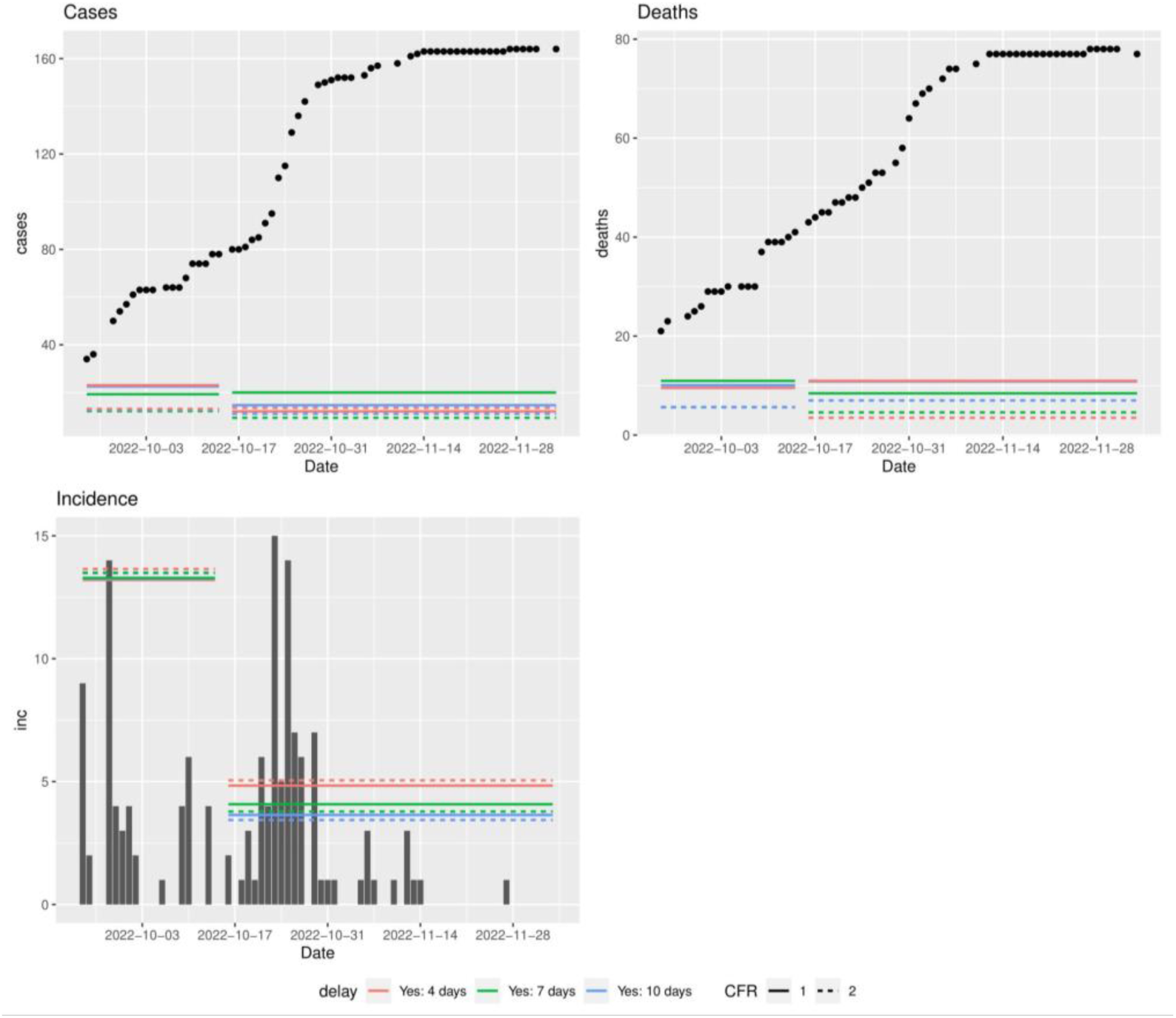
Recorded (a) cumulative case, (b) cumulative death, and (c) daily incidence counts with time-specific MSE values for ‘Two decay’ models run from 9/24 – 12/4 with diminished effects of the first intervention and one or two fatality rates.

**Table S1:**
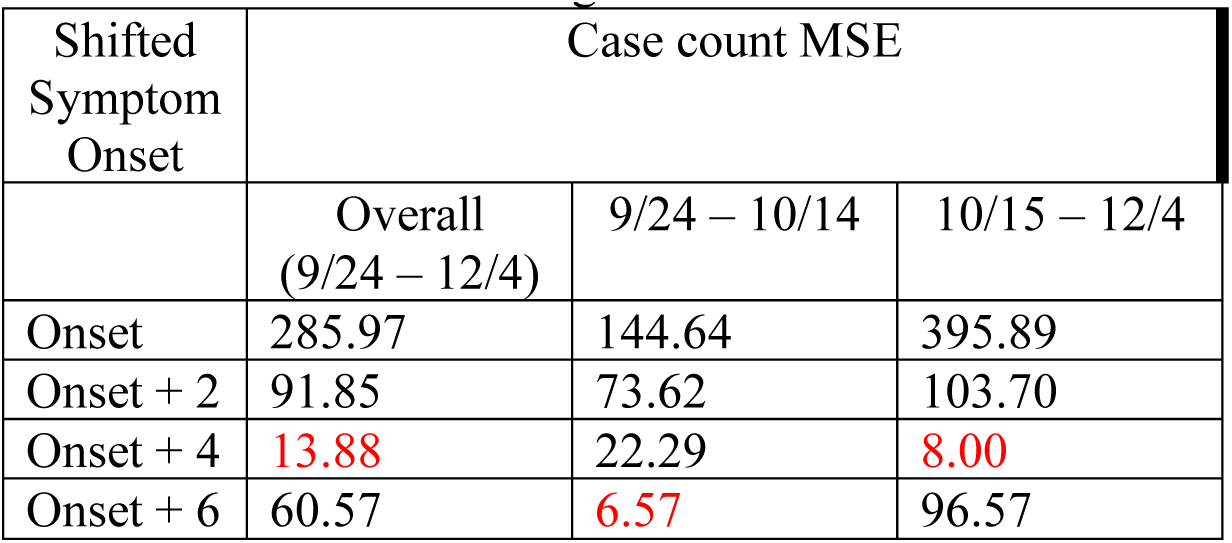
Mean Squared Error (MSE) values comparing cumulative case counts from the “as reported” dataset to those from the “MoH onset” data shifted 0, 2, 4, and 6 days forward. Values are provided for 9/24 – 10/14, 10/15 – 12/4, and the overall period (9/24-12/4), with the lowest MSE value for each date range in red.

| Shifted Symptom Onset | Case count MSE |  |  |
| --- | --- | --- | --- |
|  | Overall (9/24 – 12/4) | 9/24 – 10/14 | 10/15 – 12/4 |
| Onset | 285.97 | 144.64 | 395.89 |
| Onset + 2 | 91.85 | 73.62 | 103.70 |
| Onset + 4 | 13.88 | 22.29 | 8.00 |
| Onset + 6 | 60.57 | 6.57 | 96.57 |

**Table S2:** Mean Squared Error (MSE) values comparing cumulative case counts from the “as reported” dataset to those from the “MoH onset” data shifted 0, 2, 4, and 6 days forward. Values are provided for 9/24 – 10/14, 10/15 – 10/27, 10/28 – 12/4, and the overall period (9/24-12/4), with the lowest MSE value for each date range in red.

| Shifted Symptom Onset | Case count MSE |  |  |  |
| --- | --- | --- | --- | --- |
|  | Overall (9/24 – 12/4) | 9/24 – 10/14 | 10/15-10/27* | 10/28 – 12/4 |
| Onset | 285.97 | 144.64 | 645.55 | 3.57 |
| Onset + 2 | 91.85 | 73.62 | 170.33 | 3.75 |
| Onset + 4 | 13.88 | 22.29 | 9.50 | 5.75 |
| Onset + 6 | 60.57 | 6.57 | 165.45 | 20.80 |

\* October 27th was chosen as the place to split the data based on trends seen in Figure 1a.

**Table S3:**
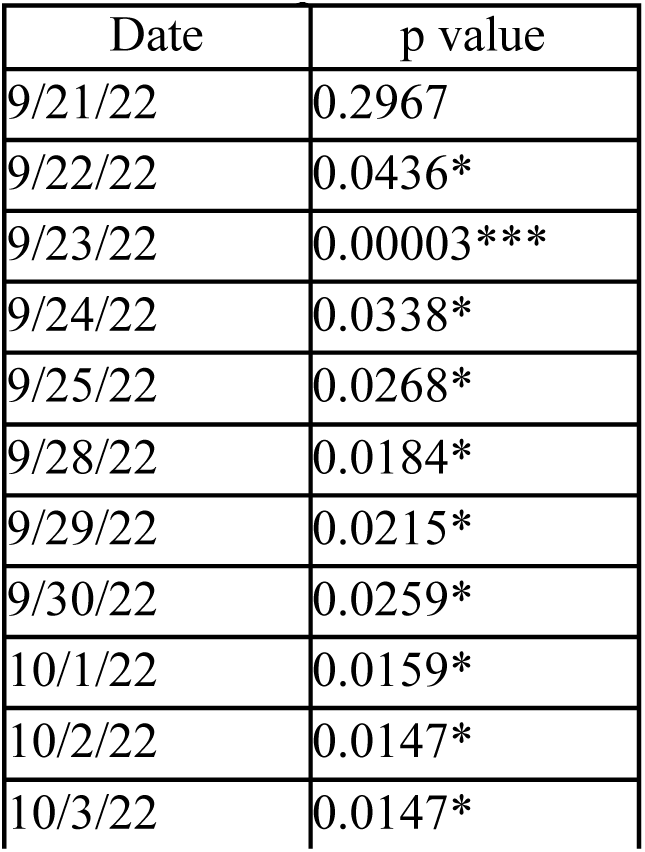
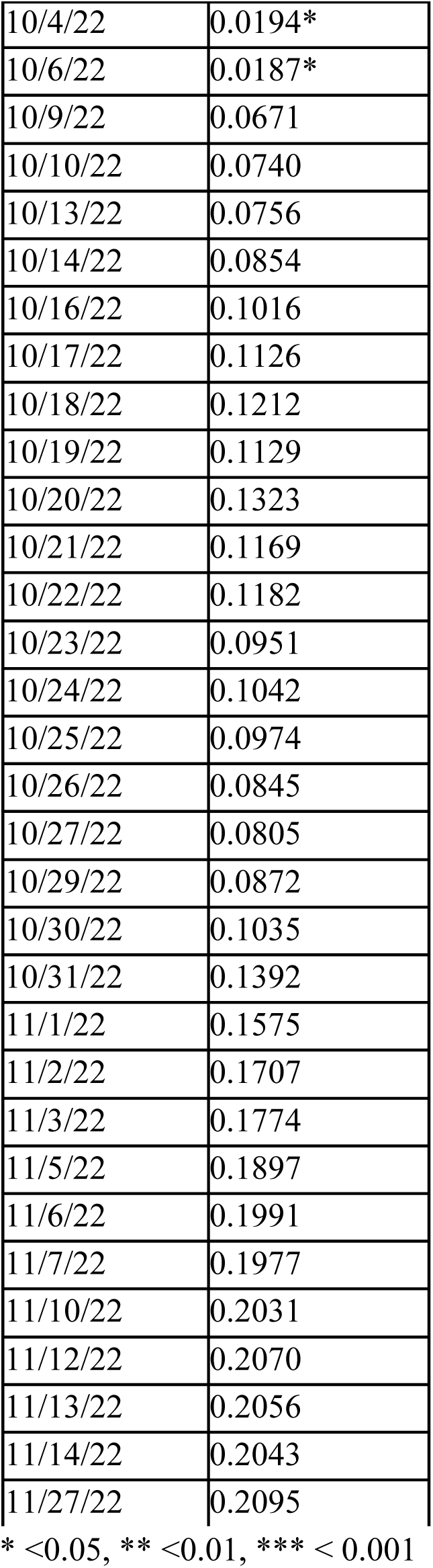
Daily Chi-squared Test of Homogeneity p-values comparing case fatality ratio (CFR) from the “as reported” and the “confirmed reported only” datasets.

